# Removing array-specific batch effects in GWAS mega-analyses by applying a two-step imputation workflow reveals new associations for thyroid volume and goiter

**DOI:** 10.1101/2024.11.21.24317711

**Authors:** M. Kamal Nasr, Eva König, Christian Fuchsberger, Sahar Ghasemi, Uwe Völker, Henry Völzke, Hans J. Grabe, Alexander Teumer

## Abstract

**Background:** Combining individual-level data in genetic association studies (mega-analyses) enhances statistical power for identifying gene-trait associations. However, batch effects from combining variants of different arrays pose a major limitation. Here, we developed a two-step imputation workflow to overcome the array type bias.

**Methods:** Genotype data of 10,647 individuals generated using five different arrays were included. Intermediate array-specific panels were generated and subsequently imputed against the 1000 Genomes Project Phase3 reference panel. Genetic principal component (PC) analysis assessed batch effects in the cohort-combined imputed data. The workflow’s performance was evaluated by comparing imputation quality r^2^ and allele frequency difference of the proposed two-step imputation to the conventional array-specific imputation as well as its matching with a whole-genome sequenced subgroup for further validation. We performed a genome-wide association study (GWAS) to test for genetic associations with goiter risk and thyroid gland volume, comparing summary statistics of both approaches.

**Results:** The proposed workflow eliminated the batch effect from the first twenty genetic PCs. The outcome of the workflow also showed high correlation with the conventional approach for allele frequencies (r^2^ > 0.99). GWAS results from the two-step imputation confirmed known associations on thyroid traits and revealed novel loci for thyroid volume (*TG, PAX8, IGFBP5, NRG1*), and one novel locus for goiter (*XKR6*), which was not statistically significant following the GWAS meta-analysis of conventional imputation.

**Conclusion:** Our imputation workflow provides high-quality imputation results without technical batch effects, fostering mega-analysis involving multiple genotyping arrays for different genetic association analysis.

## Introduction

Genome-wide association studies (GWAS) represent an agnostic approach for identifying genetic associations with common traits and diseases by testing millions of variants with continuous outcomes or between groups. The testing checks allele frequency differences between individuals in a selected population that show different representations of the trait value ^1^. GWAS analyses have been utilized in more than 5,700 studies, exploring 3,300 different traits ^1,2^. The results of these analysis enriched our knowledge about disease risk variants, as well as identifying individuals with high disease-risk profiles through risk scores for complex heritable traits ^1,3^.

The power to detect associations increases with sample size and number of variants tested ^3,4^, rare and low frequency single nucleotide variants (SNVs) are of high interest. This requires the inclusion of large sample sizes in the experimental design, which is complicated or even unrealistic for rare diseases and low number of cases in the investigated populations. One alternative approach is a meta-analysis of summary statistics from different GWAS analyses of the same trait, performed either on same or different populations ^4,5^. Another approach is to combine the individual-level data of samples from different cohorts to perform a mega-analysis ^6^.

Meanwhile, genetic imputation is a reliable method to estimate alleles of variants not directly genotyped on an array. Based on linkage disequilibrium (LD), genetic imputation can significantly increase coverage of the human genome when using different commercial genotyping arrays ^7,8^. This method is utilized as an alternative for expensive whole genome sequencing ^9^.

GWAS on imputed variants and subsequent meta-analysis are frequently utilized cost-effective approaches for conducting large GWAS^10^, however, they are also associated with technical limitations ^11^. If the sample size or number of cases in a specific cohort especially in a (nested) case-control design is low, the analysis results are less reliable because the effective number of samples is too small for the association models like linear regression, logistic regression or mixed-effects models, particularly when it comes to low-frequency variants. In addition, the analysis workload substantially increases with the number of cohorts included in a project ^12^.

Former studies have shown that both meta-analyses and mega-analyses using individual participant data are mathematically equivalent ^13^, and also comparable when using imputed genotypes ^14^. However, previous studies have mainly focused on data from cohorts genotyped on the same array type. Highlighting challenges in mega-analysis, where individual-level data from multiple cohorts genotyped on different arrays are combined.

We identified a concerning technical bias when combining the imputed data of individuals from genetically homogeneous northeastern German cohorts that were genotyped using diverse array types ^15,16^. Principal component analysis (PCA) conducted on the quality-controlled genotype data revealed variation influenced by the array type. Notably, this variation was detected upon both imputation against the haplotype reference consortium (HRC) ^17^ and the 1000 Genomes v5 (1000G) reference panels ^18^.

Here, we propose a newly developed workflow, to minimize the technical bias due to batch effect when combining genotype data from different arrays. The workflow is composed of two imputation steps. First we impute the included genotype datasets pairwise against each other and then create a panel of overlapping variants for each imputation outcome. Finally, we impute the generated intermediate panel against one of the commonly available large panels. We evaluate the outcome of the new workflow in comparison to conventional imputation approaches. In an application example, we conducted a GWAS analysis on thyroid gland volume and goiter, identifying new associations with goiter while demonstrating the robustness of our imputation workflow by validating known associations.

## Materials and Methods

### Workflow design overview

In this project, three different approaches for genetic imputation have been conducted. The first proposed approach is a new two-step imputation process. The second and third approaches are conventional single-step imputation for comparison with the newly proposed method. We modified the third approach to be a single-step imputation using only the intersecting variants genotyped on all included array types. The genotype data that were used for imputation were the same in both imputation approaches. We used data from five different arrays (Affymetrix SNP 6.0, Affymetrix Axiom [Thermo Fisher Scientific, Santa Clara, CA, USA], and Illumina Omni 2.5, Illumina GSA, and Illumina PsychArray [Illumina, Inc., San Diego, CA, USA]) obtained from samples of the German GANI_MED (n= 2410), SHIP-START (n=4070) and SHIP-TREND (n= 4119) (Figure 1) ^15,16^. The individuals genotyped on the Affymetrix Axiom array (n= 48) were a subset of the Affymetrix SNP 6.0 samples, whereas all other individuals were genotyped only once. Detailed information regarding the included datasets, sample sizes, and genotype arrays are provided in Figure 1 and the Supplementary Methods. For evaluation of the imputation performance, a whole-genome sequenced subset of 192 individuals from SHIP-TREND has been used for genotype matching concordance checking.

**Figure 1.**
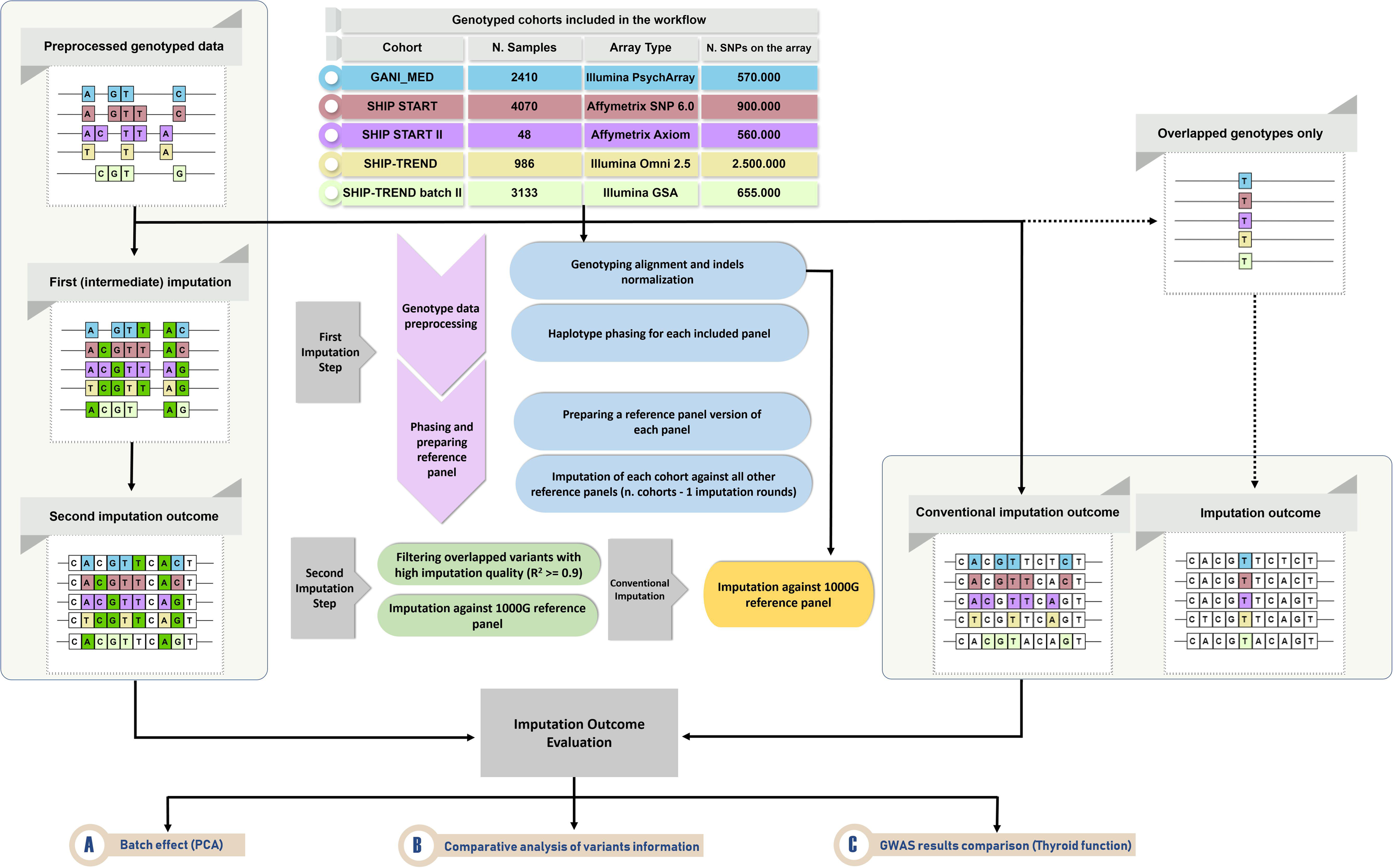
Overview of the designed workflow. Variants colored in dark green represent the variants imputed in the intermediate imputation phase (first imputation), while variants colored in white are imputed from general 1000G reference panel in both two-step and conventional imputation.

### Data pre-processing and phasing

Genotype quality control was performed using PLINK ^19^ as reported previously ^20,21^. Briefly, arrays with genotype call rate <94%, as well as variants with missing call rate >5%, Hardy-Weinberg equilibrium p-value <10^-4^, and monomorphic SNVs and singletons were excluded. All the included genotype datasets were aligned to reference genome build (GRCh37) using BCFtools software followed by retaining only sites with at least one alternative allele ^22^. Haplotype phasing preceding the first imputation round was performed for each genotype dataset using Eagle2 (5 phasing iterations and number of conditioning haplotypes = 10^4^) and estimated to hg19 mapping ^23^. No external reference panel was used for phasing.

### Two-step imputation

In the first step, the intermediate imputation, each phased panel was used as a reference panel after compression with the minimac4 imputation software ^24^. Each of the five pre-processed genotype arrays was imputed pairwise against the other four, yielding twenty imputed datasets. Using an R scripted algorithm with VCFtools and BCFtools for variants filtration ^22^, each of these imputed variant sets were subset to the variants overlapping between all generated panels with high imputation quality (R^2^ >= 0.9), selecting the source panel with the highest imputation quality score for each variant. The outcome of the first-step imputation was one VCF file for each cohort dataset, which was then used as input for the second imputation step against 1000G reference panel using the Michigan imputation server ^24^. Eagle2 was selected for phasing without (additional) imputation quality filters applied at this stage of the imputation.

### Conventional imputation

To validate the outcome of the proposed workflow, we imputed the genotype data of each array directly against 1000G reference panel using the same parameters and quality control procedures as described in the two-step imputation.

### Conventional imputation with overlapping genotypes

To evaluate a simple approach for removing the array type specific batch effect, we ran a single-step imputation by restricting the input variants to those assessed on all array types. This approach was tested for three (Affymetrix SNP 6.0, Illumina Omni 2.5 and Illumina GSA) and all five array types. We compared the imputation quality of these imputations to the conventional and the two-step imputation exemplarily for the SHIP-TREND.

### Batch effect assessment

Technical bias was assessed by estimating genetic principal components of the imputed and quality-controlled genotype data. Quality control filters included missing variant call rates above 5%, Hardy-Weinberg equilibrium p-values less than 10^-4^, minor allele frequency (MAF) less than 1%, correlated variants were removed by LD pruning with a window size of 50, step size of 5 SNVs and R^2^ threshold of 0.2. Genetic principal components, along with their explained variances were compared between two-step imputation and conventional imputation. To check the robustness of the two-step imputation approach for rare variants, we further ran PCA on rare variants (MAF < 1%). All PCAs are performed using PLINK 2.0 software ^19^. Principal components (PC) of the first twenty components were plotted with ggplot2 package of R software.

### Evaluation of imputation performance

Genotype data attributes, including differences in MAF and imputation quality R^2^ measure between the two-step and conventional imputation were compared for each included cohort after stratification by minor allele frequency. We additionally checked the concordance (number of matching genotypes/total number of genotypes ^25^) of the imputed best-guess (hard call) genotypes with their whole-genome sequencing data of a subset of 192 individuals from SHIP-TREND (Omni 2.5) for which whole genome sequencing (WGS) data was also available.

### GWAS and results comparison

We evaluated the results of GWAS analyses using genotype dosage information from both two-step and conventional imputation approaches using thyroid gland volume and goiter risk as outcomes as described before ^26^, and summarized in the Supplementary Methods.

All GWAS were conducted with the EPACTS 3.3.2 software using dosages ^27^. Inverse-variance weighted meta-analysis of the single cohort GWAS summary statistics based on the conventional imputation approach was done using the METAL software ^4^. We compared the summary statistics p-values, effect estimates and its standard errors. A detailed description of the GWAS analysis plan is available in the Supplementary Methods. Independent (lead) variants associated with thyroid traits were identified using the clumping function of PLINK with a threshold of p < 5 x 10^-8^, r^2^ > 0.01 and 1 Mb distance, and compared across the imputation approaches. For biological validation of the findings, lead variants were investigated by checking their association with thyrotropin to support their biological plausibility ^20^.

## Results

Genotype datasets of 10,647 individuals genotyped on five different array types were included in the workflow (Figure 1). After the intermediate imputation, 1,942,499 high-quality overlapping autosomal variants were generated for each of the included five datasets, and subsequently used for the second imputation step against the 1000G reference panel. In contrast, only 58,091 autosomal genotyped variants were available as imputation input in the three cohorts using the conventional imputation approach with overlapping variants. This number further dropped to 12,265 variants when including the Affymetrix Axiom and Illumina PsychArrays. Supplementary Table 1 provides the distribution of these variants for each chromosome in the two-step imputation, the conventional imputation and conventional imputation using overlapped variants.

### Batch effect investigation (PCA analysis)

For the conventional imputation, 939,802 variants with a total genotyping rate of 0.988 were included in the PCA analysis, with 2,314,205 variants removed due to missing genotype threshold, 9,118 variants due to Hardy-Weinberg equilibrium, and 38,008,458 variants due to the MAF threshold. Projecting the first two principle components of the imputed genotypes clustered the samples by array types with a combined explained variance of 0.242 (Figure 2a, b, Supplementary Figure 1a).

**Figure 2.**
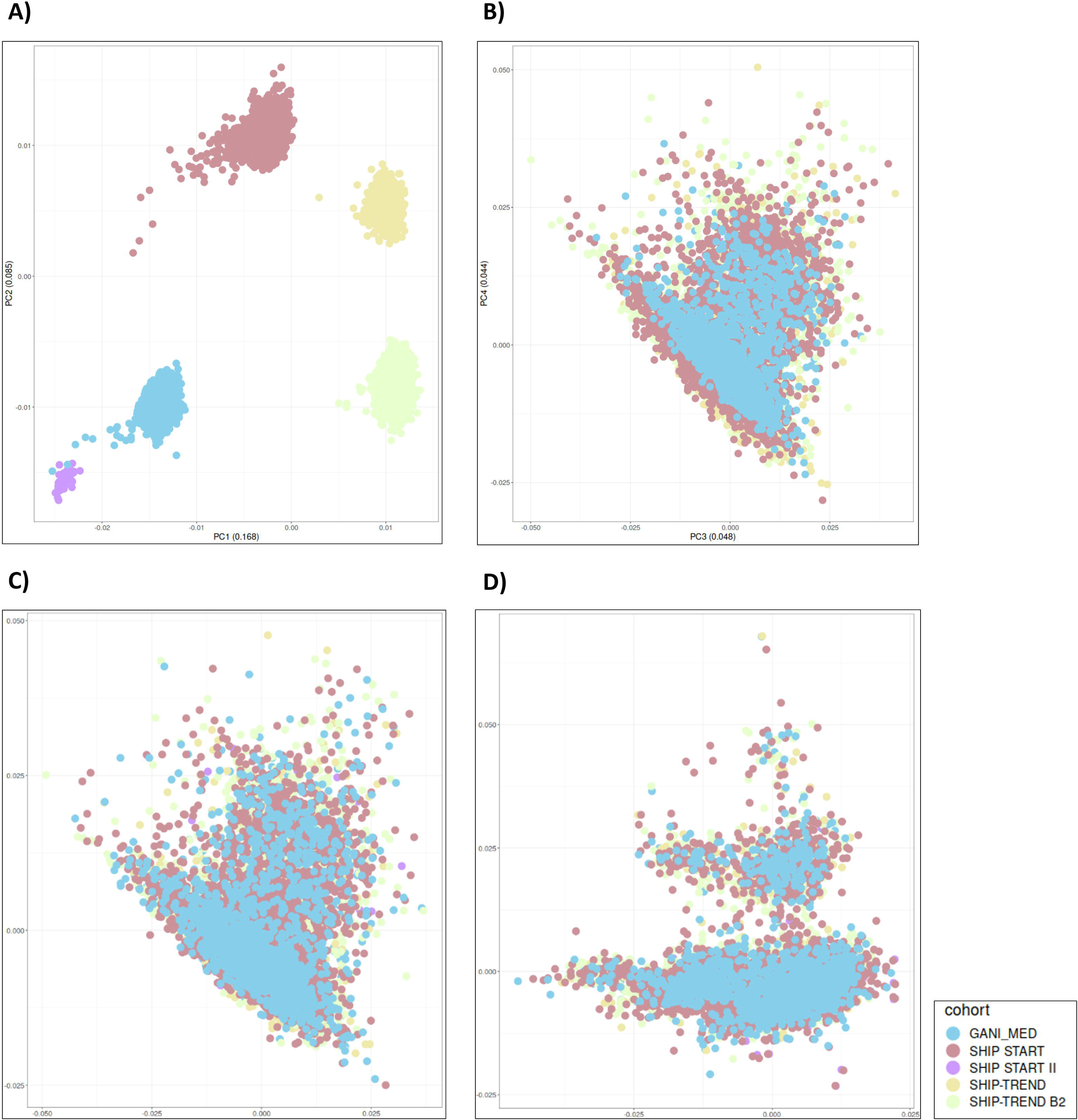
Genetic PCA of the included cohorts in the workflow. The samples are colored by the cohorts with their unique array type. Panel A and B show the first four genetic components of the conventional imputation approach (variance explained for the components combined is 0.34). Panels C and D show the first four components of the proposed approach (variance explained for the components combined is 0.17).

The two-step imputation included 1,360,834 variants with a total genotyping rate of 0.989 in the PCA analysis after removing 725,008 variants due to missing genotype threshold, 5,363 variants due to Hardy-Weinberg equilibrium, and 37,753,448 variants due to the MAF threshold. Using this approach, the first 20 principal components of the imputed genotypes did not show any array type specific clusters (Figure 2c, d, Supplementary Figure 1b). PCA restricted to the rare genotypes imputed with the two-step approach did not show clustering by the array type (Supplementary Figure 2).

The conventional imputation using only overlapping variants also removed the batch effect (Supplementary Figure 3), but led to a drastic decrease in the quality of the imputed variants for all allele frequency groups in comparison to the other approaches (Supplementary Figure 4). Median R^2^ of the overlapping variants was 0.011, while for two-step imputation and conventional imputation using all variants was 0.993 for variants with MAF > 0.05, which was the main reason for not considering this approach any further in the performance evaluation and GWAS analysis, focusing only on the two-step imputation against conventional imputation using all variants.

### Two-step imputation outcome parameters comparison with conventional imputation

Our newly developed two-step imputation approach showed higher overall quality for the imputation outcome of rarer variants (MAF < 0.01) than the conventional imputation (Figure 3, Supplementary Figure 5). For less-common variants with MAF between 0.04 and 0.05, the median R^2^ increased by 0.19 when using the two-step imputation approach for the GANI_MED Illumina PsychArray. It also shows comparable allele frequencies to the conventional approach, judging by the absolute difference in allele frequencies for each imputed variant in both approaches (Figure 4). Detailed statistics about median R^2^ and MAF differences are presented in Supplementary Table 2.

**Figure 3.**
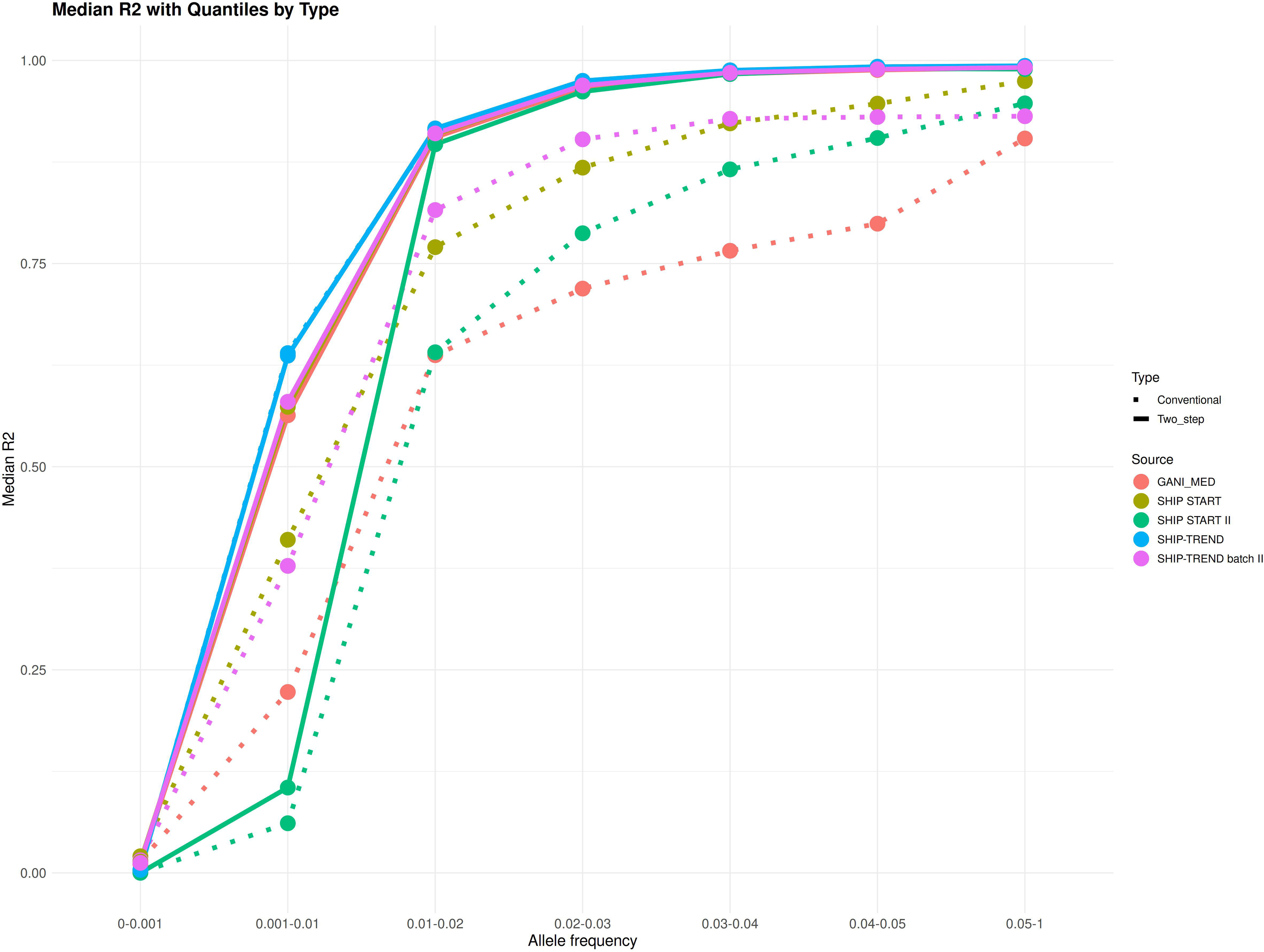
Median R2 of the imputation outcomes of genotype data of the included cohorts, using conventional (dotted) and two-step imputation (full line).

**Figure 4.**
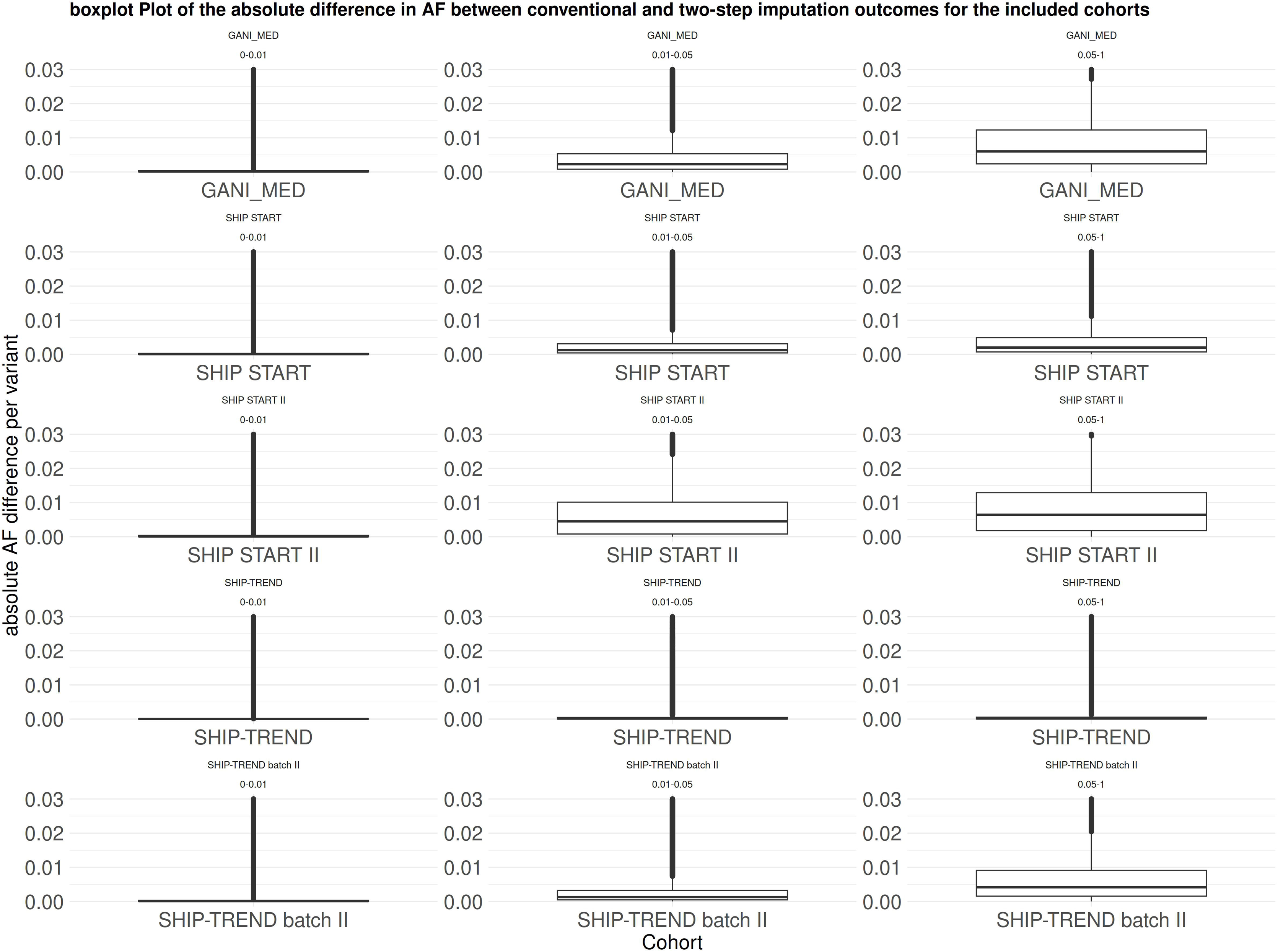
Boxplots grid of the absolute difference in allele frequency (AF) between conventional and two-step imputation outcomes for the included cohorts. Each column represents an allele frequency group (rare, low, common) and each row represents one of the included cohorts.

Hard call genotype concordance results in the subgroup of SHIP-TREND with matching WGS data showed strong correlation between two-step imputation and sequenced genotypes. The Pearson correlation coefficients with the homozygous reference, heterozygous, and homozygous alternative genotypes were 0.996, 0.981, and 0.979, respectively (Table 1, Supplementary Figure 6).

**Table 1.** Genotype concordance (number of matching genotypes/total number of genotypes) of the hard call imputed genotypes with sequenced data for homozygous reference (HomRef), homozygous alternative (HomAlt), and heterozygous (Het) calls, all variants followed by stratification by minor allele frequency (MAF) group of the imputed data. (n) represents number of variants represented per group in both imputed and sequenced genotypes.

| Group | All variants |  | MAF 0.05-0.5 |  | MAF 0.01-0.05 |  | MAF 0.0-0.01 |  |
| --- | --- | --- | --- | --- | --- | --- | --- | --- |
|  | Conventional | Two-step | Conventional | Two-step | Conventional | Two-step | Conventional | Two-step |
| HomRef | 0.9963<br>(618,453,288) | 0.9962<br>(613,997,680) | 0.9913<br>(224,034,629) | 0.9910<br>(222,769,137) | 0.9959<br>(171,621,821) | 0.9988<br>(183,771,150) | 0.9993<br>(222,756,838) | 0.9994<br>(207,703,823) |
| Het | 0.9814<br>(116,274,486) | 0.981<br>(116,144,961) | 0.9897<br>(105,647,496) | 0.9893<br>(105,617,755) | 0.9424<br>(8,530,966) | 0.9418<br>(8,903,479) | 0.7875<br>(2,139,474) | 0.7525<br>(1,695,994) |
| HomAlt | 0.9783<br>(21,587,792) | 0.9787<br>(21,408,323) | 0.9802<br>(21,463,336) | 0.9807<br>(21,290,637) | 0.7979<br>(128,442) | 0.7862<br>(124,359) | 0.2512<br>(5,149) | 0.1389<br>(2,369) |

### GWAS on thyroid traits

A total number of 6,894 individuals from SHIP-START and SHIP-TREND (both array types) with thyroid measurement information were included in the GWAS analysis, all participants were of European ancestry. Detailed information about cohort characteristics is presented in Table 2. The estimated genomic control for all conducted GWAS analysis showed no signs of inflation, with a minimum and maximum __GC_ = 1.001 and 1.038, respectively (Supplementary Figure 7).

**Table 2.** Cohort characteristics for GWAS analysis of thyroid volume and goiter risk. BSA: body surface area.

| Cohort | N | Age<br>(mean ± SD) | Sex | BSA<br>(mean ± SD) | Current<br>smoker | Log thyroid volume<br>(mean ± SD) | Goiter<br>cases(%) |
| --- | --- | --- | --- | --- | --- | --- | --- |
| SHIP START | 3,611 | 49.1 (16.3) | 47.4% Male,<br>52.6% Female | 1.9 (0.2) | 31.6% | 2.9 (0.5) | 1322 (36.6%) |
| SHIP-TREND | 784 | 49.2 (13.9) | 51.5% Male,<br>48.5% Female | 1.9 (0.2) | 22.2% | 2.9 (0.4) | 253 (32.3%) |
| SHIP-TREND<br>batch II | 2,499 | 51.3 (16) | 56.9% Male,<br>43.1% Female | 1.9 (0.2) | 29.8% | 2.9 (0.4) | 737 (29.5%) |

Meta-analysis of the GWAS analysis on goiter risk using conventionally imputed genotypes revealed four significantly associated loci (p < 5 x 10^-8^). Confirming the results from previously conducted GWAS analysis^26^. Two of the loci are located at the *CAPZB* region in chromosome 1, the other two at the *FAM227B* and *MAFTRR* regions on chromosome 15 and 16, respectively (Figure 5b). However, the GWAS analysis of the combined two-step imputed genotype data revealed another associated locus at the *XKR6* region on chromosome 8 (Figure 5c), which did not attain statistical significance using the conventional meta-analysis approach (Figure 5a).

**Figure 5.**
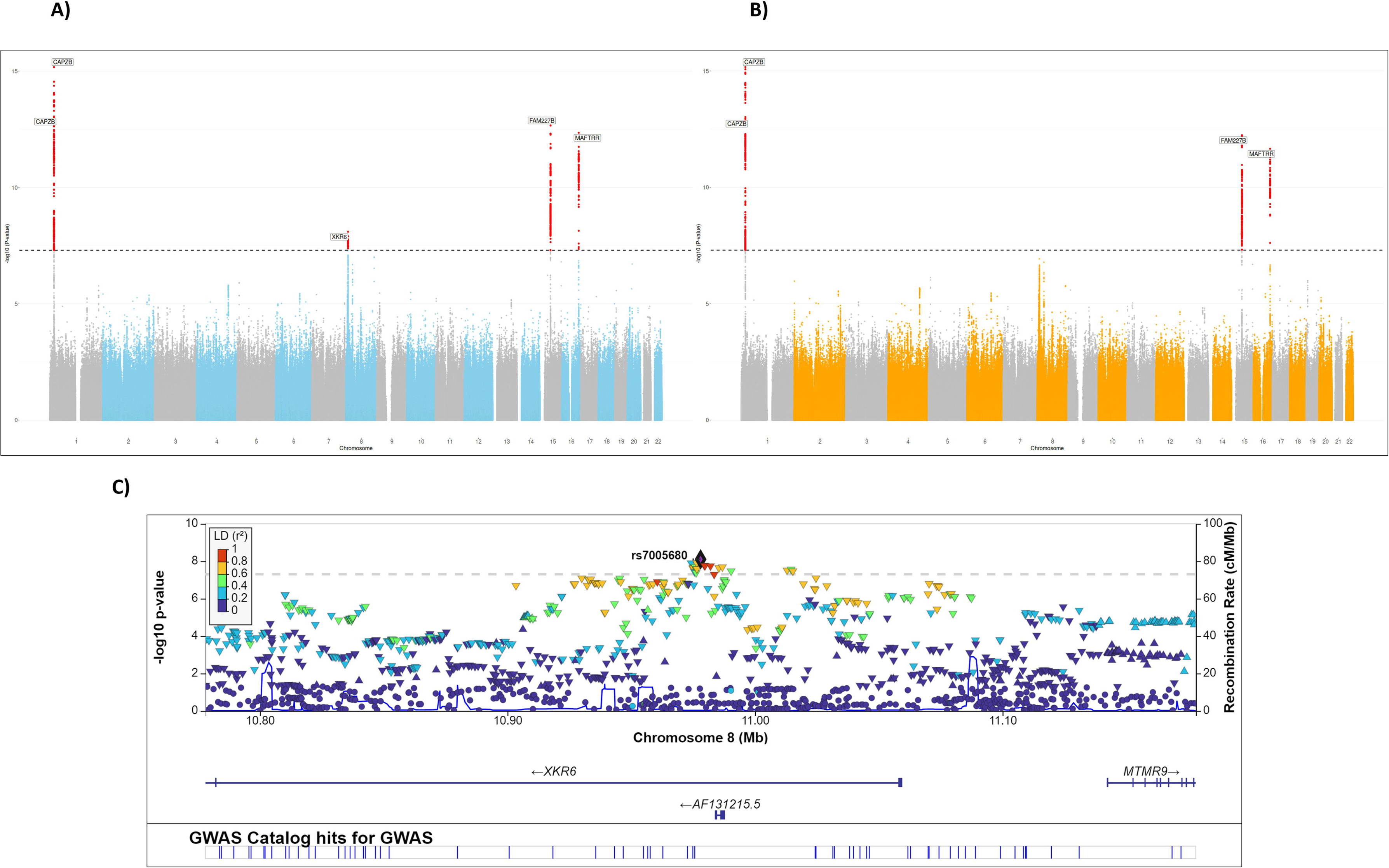
Manhattan plot of the GWAS analysis of goiter risk using combined two-step imputation genotypes (A) and conventional imputation and meta-analysis approach (B). Variants are plotted on the x axis and –log_10_ p-values of the association testing on the y axis. Associations significant after correction for multiple testing (p < 5 ×10^-8^) are colored in red. Regional association results and recombination rates for the *XKR6* gene from two-step imputation GWAS are presented in part C, −log_10_ p-values (y-axis) of the single nucleotide variants according to their chromosomal positions (x-axis) with lead variant (rs7005680) is shown as a purple diamond.

The conventional approach of the GWAS meta-analysis of thyroid volume revealed four novel associations at the *PAX8* region on chromosome 2, at *IGFBP5*, *NRG1* and *TG* (Figure 6b,c), and confirmed all known associations with this trait^20^. Genome-wide significance for these regions were also obtained with the GWAS using the combined two-step imputed genotype data, with the exception of *NRG1* (p-value = 5.22 x 10^-8^). (Figure 6a). Except the *PAX8* region, all associated loci were also associated with thyroxin in recently published multi-trait GWAS meta-analysis analysis for thyroid function ^20^.

**Figure 6.**
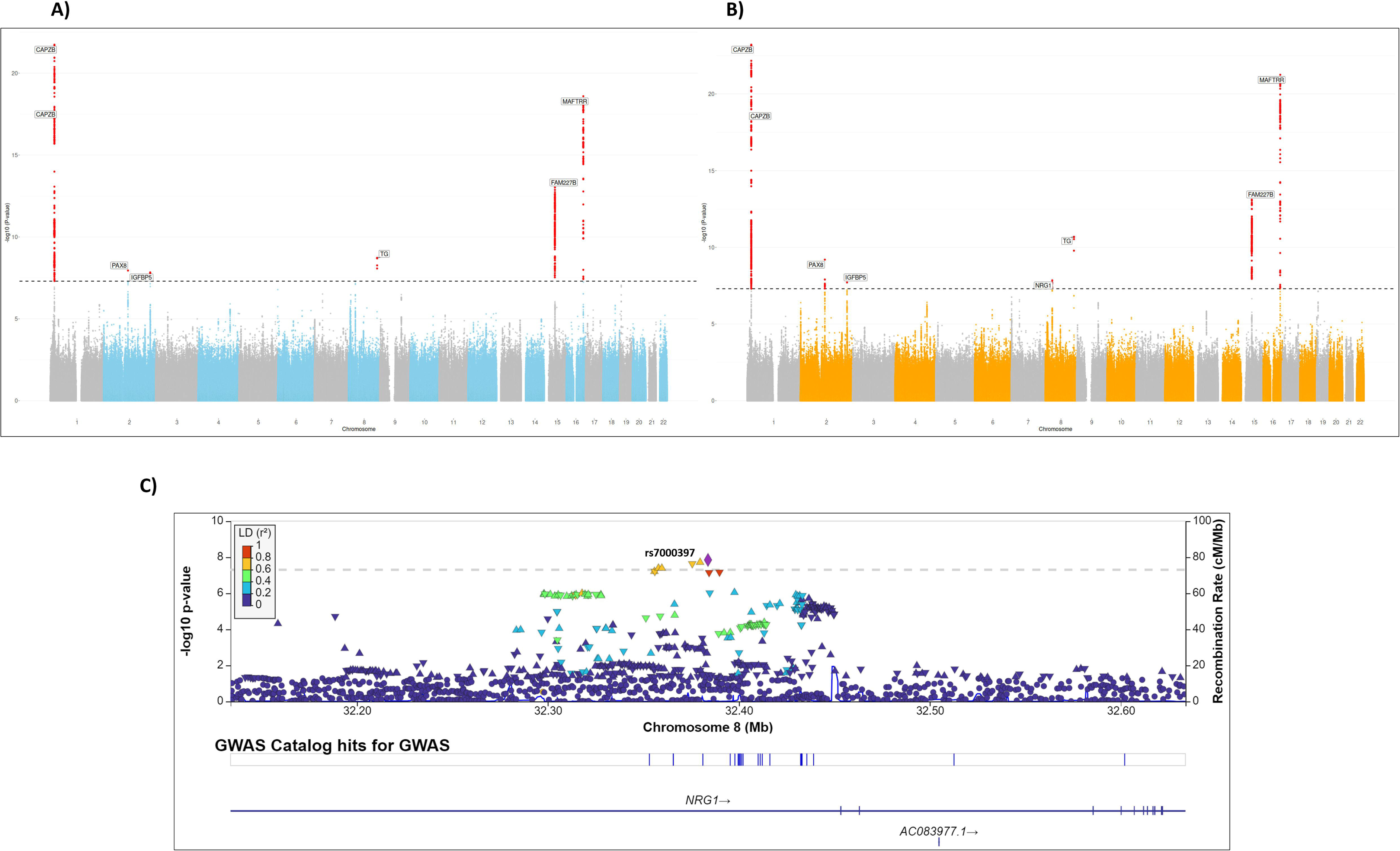
Manhattan plot of the GWAS analysis of log thyroid volume using combined two-step imputation genotypes (A) and conventional imputation and meta-analysis approach (B). Variants are plotted on the x axis and –log_10_ p-values of the association testing on the y axis. Associations significant after correction for multiple testing (p < 5 ×10^-8^) are colored in red. Regional association results and recombination rates for the *NRG1* gene from conventional imputation GWAS are presented in part C, –log_10_ p-values (y-axis) of the single nucleotide variants according to their chromosomal positions (x-axis) with lead variant (rs7000397) is shown as a purple diamond.

Table 3 summarizes the results of the SNVs with the strongest association of both GWAS approaches and traits. The significant GWAS results of both approaches were comparable, judging by the magnitude of the estimates. However, the standard errors of the GWAS results obtained from the two-step imputed genotypes where generally lower, where the natural log p-values were slightly higher in the linear regression and lower in the logistic regression based analyses compared to the conventional imputation and subsequent meta-analysis. (Supplementary Figures 8 and 9).

**Table 3.**
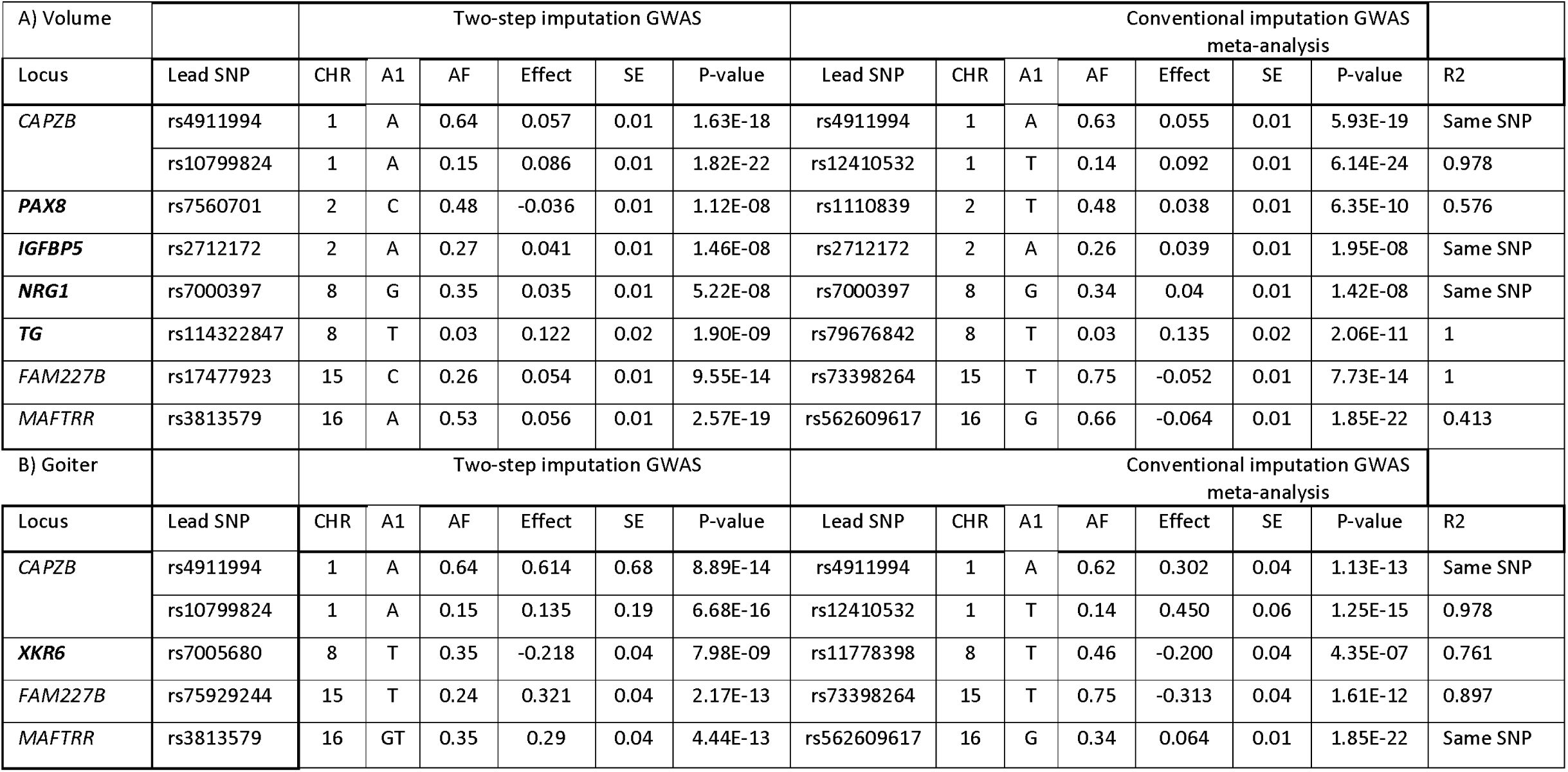
Loci lead SNVs with the strongest association with thyroid volume (A), and goiter risk (B) in GWAS analysis using both two-step imputed and conventionally imputed genotypes. Novel loci are marked in bold. A1: coded allele, AF: frequency of coded allele, SE: standard errors, R2: linkage disequilibrium in the top SNVs of the same locus in both imputation outcomes

| A) Volume |  | Two-step imputation GWAS |  |  |  |  |  | Conventional imputation GWAS meta-analysis |  |  |  |  |  |  |  |
| --- | --- | --- | --- | --- | --- | --- | --- | --- | --- | --- | --- | --- | --- | --- | --- |
| Locus | Lead SNP | CHR | A1 | AF | Effect | SE | P-value | Lead SNP | CHR | A1 | AF | Effect | SE | P-value | R2 |
| CAPZB | rs4911994 | 1 | A | 0.64 | 0.057 | 0.01 | 1.63E-18 | rs4911994 | 1 | A | 0.63 | 0.055 | 0.01 | 5.93E-19 | Same SNP |
|  | rs10799824 | 1 | A | 0.15 | 0.086 | 0.01 | 1.82E-22 | rs12410532 | 1 | T | 0.14 | 0.092 | 0.01 | 6.14E-24 | 0.978 |
| PAX8 | rs7560701 | 2 | C | 0.48 | -0.036 | 0.01 | 1.12E-08 | rs1110839 | 2 | T | 0.48 | 0.038 | 0.01 | 6.35E-10 | 0.576 |
| IGFBP5 | rs2712172 | 2 | A | 0.27 | 0.041 | 0.01 | 1.46E-08 | rs2712172 | 2 | A | 0.26 | 0.039 | 0.01 | 1.95E-08 | Same SNP |
| NRG1 | rs7000397 | 8 | G | 0.35 | 0.035 | 0.01 | 5.22E-08 | rs7000397 | 8 | G | 0.34 | 0.04 | 0.01 | 1.42E-08 | Same SNP |
| TG | rs114322847 | 8 | T | 0.03 | 0.122 | 0.02 | 1.90E-09 | rs79676842 | 8 | T | 0.03 | 0.135 | 0.02 | 2.06E-11 | 1 |
| FAM227B | rs17477923 | 15 | C | 0.26 | 0.054 | 0.01 | 9.55E-14 | rs73398264 | 15 | T | 0.75 | -0.052 | 0.01 | 7.73E-14 | 1 |
| MAFTRR | rs3813579 | 16 | A | 0.53 | 0.056 | 0.01 | 2.57E-19 | rs562609617 | 16 | G | 0.66 | -0.064 | 0.01 | 1.85E-22 | 0.413 |
| B) Goiter |  | Two-step imputation GWAS |  |  |  |  |  | Conventional imputation GWAS meta-analysis |  |  |  |  |  |  |  |
| Locus | Lead SNP | CHR | A1 | AF | Effect | SE | P-value | Lead SNP | CHR | A1 | AF | Effect | SE | P-value | R2 |
| CAPZB | rs4911994 | 1 | A | 0.64 | 0.614 | 0.68 | 8.89E-14 | rs4911994 | 1 | A | 0.62 | 0.302 | 0.04 | 1.13E-13 | Same SNP |
|  | rs10799824 | 1 | A | 0.15 | 0.135 | 0.19 | 6.68E-16 | rs12410532 | 1 | T | 0.14 | 0.450 | 0.06 | 1.25E-15 | 0.978 |
| XKR6 | rs7005680 | 8 | T | 0.35 | -0.218 | 0.04 | 7.98E-09 | rs11778398 | 8 | T | 0.46 | -0.200 | 0.04 | 4.35E-07 | 0.761 |
| FAM227B | rs75929244 | 15 | T | 0.24 | 0.321 | 0.04 | 2.17E-13 | rs73398264 | 15 | T | 0.75 | -0.313 | 0.04 | 1.61E-12 | 0.897 |
| MAFTRR | rs3813579 | 16 | GT | 0.35 | 0.29 | 0.04 | 4.44E-13 | rs562609617 | 16 | G | 0.34 | 0.064 | 0.01 | 1.85E-22 | Same SNP |

## Discussion

Genome-wide association studies have helped identify genetic factors for many traits and diseases. Since the 1960s, collecting genotype samples from diverse populations has grown in importance ^28,29^. Imputation techniques now address gaps in whole-genome sequencing ^30^. However, variations in these methods across cohorts reduce the accuracy of genetic analyses due to biases from differing array technologies ^31,32^. For instance, in our genotyped samples where we compared the allele frequencies of the SHIP-TREND subgroup genotyped using Illumina Omnia 2.5 with the corresponding whole genome sequencing variants (Supplementary Figure 10), this array-specific variation in the allele frequency, both in common and rare variants, can affect LD estimation in haplotype phasing and genotype imputation. Our newly developed imputation method addresses the additional variation induced by including multiple array types. By forming an intermediate panel of high-quality variants, the method enables high imputation accuracy while removing the array type induced batch effects.

The developed workflow was inspired by the use of only overlapping variants in the included cohorts (Supplementary Table 1), which eliminated the observed bias, yet led to a significant decrease in the imputation quality due to removal of informative tag SNVs (Supplementary Figure 3), the influence of informative SNVs on imputation quality has been shown in previous research work ^33^. Based on that, we introduced an intermediate imputation step to generate a panel that can retain the same genotype information of the included arrays and thus preserving its LD structure.

Selecting an appropriate threshold for the imputation R^2^ (≥ 0.9) of the overlapping variants in the intermediate panel was essential for having reliable genotype information upon imputing against 1000G panel. The value of the threshold was decided following the output of several imputations using different thresholds. We aimed to use the highest possible value for imputation quality R^2^ without affecting haplotype phasing results due to removing too many variants. Supplementary Table 1 shows the number of included variants per chromosome when the R^2^ threshold was adjusted to 0.8 as well as 0.9. Both approaches led to similar imputation outputs as seen after plotting genetic PCs of the imputation outcomes (Figure 2 and Supplementary Figure 11).

To evaluate the existence of the array type bias, we used genetic PCs to capture the main variance in the allele frequencies, which can be seen as the influence of the array type after projecting the PCs of the eigenvectors and evaluate the homogeneity of the projected points on PC axis ^34^. The imputation following the developed two-step approach showed its capability to overcome the array type differentiation whenever existing in the first twenty components, compared to conventional imputation outcomes where a clear clustering effect by the array type was observed (Figure 2).

To test the performance of the imputation outcomes, we focused on comparing allele frequency and imputation quality parameters of the cohort genotypes. These parameters are particularly relevant for conducting trait-association studies. The developed imputation flow showed strong matching of the allele frequency, represented by minimized difference in allele frequency for each variants between the imputation approaches (Figure 4 and Supplementary Table 2). The developed imputation workflow was successful in providing more reliable genetic predictions for imputed rarer alleles (Figure 3). Moreover, a strong correlation in the median R^2^ of the two imputation approaches using the SHIP-TREND Omni 2.5 arrays was observed in comparison to other cohorts. This could be due to the high variant coverage of the array used for genotyping the cohort’s samples. Further analysis of the allele frequency variance for each variant from both imputation outcomes of this specific cohort shows the strong correlation in the autosomal allele frequency (correlation r^2^ >0.999) (Supplementary Figure 12). Nevertheless, SHIP-TREND imputed genotypes showed strong concordance with the corresponding genotyped variants in the whole-genome sequenced subgroup, indicating the representation of well estimated genotypes in both rarer and common variants (Table 1 and Supplementary Table 3). Using the allele frequencies of the WGS subgroup as a gold standard for comparison of the imputation results is somewhat misleading because differences in the genotype frequencies exist already between genotyped variants on the arrays and the WGS (Supplementary Figure 10).

The aim of conducting GWAS as part of the imputation workflow evaluation is to compare regression analyses results of the combined two-step imputed datasets to the conventional inverse-variance meta-analysis of the same samples. Goiter risk and thyroid volume are both suitable traits for our evaluation as they represent different trait datatypes with a true positive genetic association in SHIP ^26^. The results of GWAS analysis for the two-step imputed data showed two key advantages. First, we were able to identify a new association in the *XKR6* gene region with goiter risk as a dichotomous trait which did not reach statistical significance in the conventional meta-analysis. Variants in this locus were also associated with thyroxin in a former GWAS, making this association biologically reasonable ^20^. A slightly better performance of the mega-analysis vs. a meta-analysis for a logistic regression GWAS is in line with the results of a former study ^14^. The second point was its consistency with conventional imputation GWAS meta-analysis in revealing a novel locus associated with thyroid gland volume (*PAX8*). *PAX8* is a paired box family gene member that was found to be associated with the development of thyroid gland in embryonic development, and its transcription is a diagnostic marker for anaplastic thyroid carcinoma ^35–37^, and thus represents also a plausible association with thyroid volume in adults. All other significant GWAS findings represent also plausible true positive associations as they confirmed former findings of these traits (including replication in an independent cohort)^26^, or were associated with thyroid function^20^.

The comparison of the estimates and standard errors of both GWAS approaches did not show signs of p-value inflation. However, it showed that the GWAS of the two-step imputation data had a slight decrease in standard errors, in comparison to the meta-analysis outcomes.

Besides its role in discovering more hits in case-control GWAS analysis, our developed two-step imputation workflow is not restricted to a specific array, the inclusion of different arrays with different sequencing technologies in the workflow has proven its robustness in overcoming the array bias regardless of the number or the type of the array. It is also applicable for other generalized imputation panels like TOPMed or HRC reference panels ^24^. These two strengths shall enable the utilization of combined genotype information for better understanding of rare diseases or genetic associations in populations that are represented in small sample sizes.

Although the array type specific batch effect in GWAS mega-analyses might be reduced in specific scenarios by adjusting for genetic PCs, such a correction will not be possible in all analyses. Such analyses include the ones using polygenic scores. Our two-step imputation provides a powerful solution for combining datasets while reducing technical bias also for analyses of polygenic scores.

Comparing the imputation outcomes to the whole genome-sequenced data had limitations for evaluating the proposed workflow, either due to the relatively small sample size of individuals who underwent WGS (n = 192 after QC) and its exclusivity for one of the included five cohorts. Although the total sample size provided us the possibility of analysing low frequency alleles, it was likely too small for evaluating the performance of the different approaches for very rare alleles. However, our two-step imputation approach seems to be on average superior to the classical imputation with regard to the imputation quality measure (Figure 3), while the difference in allele frequency is small particularly for rarer variants (Figure 4 and Supplementary Table 2). As indicated also in these results, the average difference in allele frequency seems to depend also on the density and design of the underlying genotyping array.

While the imputed genotypes generated from the proposed workflow will be utilized for identifying novel genetic associations in SHIP and GANI-MED, it will be interesting to evaluate the impact of the developed workflow on other cohorts, especially those comprising diverse ancestries. Thus far, all the included cohorts are of European ancestry from the North-East of Germany, highlighting the need to test the workflow’s efficacy on genotyped cohorts from other or multiple genetic ancestries. Such evaluations will help to determine the generalizability and robustness of the imputation method across different genetic backgrounds, ultimately enhancing its utility in global genomic research.

In conclusion, our developed two-step imputation workflow aims to overcome the array type bias, by creating an intermediate panel of high-quality overlapping imputed variants. This approach enables the conduction of mega-analysis by combining genotype information from different arrays without inducing a technical array type effect. Our workflow will increase statistical power for conducting large-scale GWAS mega-analyses and other genetic analyses like polygenic risk score calculations, playing an important role in genetic research and its application in individualized medicine.

## Acknowledgements

SHIP is part of the Community Medicine Research net of the University of Greifswald, Germany, which is funded by the Federal Ministry of Education and Research (grants no. 01ZZ9603, 01ZZ0103, and 01ZZ0403), the Ministry of Cultural Affairs as well as the Social Ministry of the Federal State of Mecklenburg-West Pomerania, and the network ‘Greifswald Approach to Individualized Medicine (GANI_MED)’ funded by the Federal Ministry of Education and Research (grant 03IS2061A). Genome-wide data have been partly supported by the Federal Ministry of Education and Research (grant no. 03ZIK012) and a joint grant from Siemens Healthineers, Erlangen, Germany and the Federal State of Mecklenburg-West Pomerania. The project is funded by the Deutsche Forschungsgemeinschaft (DFG, German Research Foundation) – 455978266 (A.T.). The authors are grateful to Linda Garvert for testing and helpful feedback on the two-step imputation workflow.

## Author Contributions

Project design and supervision: A.T. Analyses: M.K.N., E.K., S.G.

Interpretation of the results: M.K.N., A.T., E.K., C.F. Drafting of the manuscript: M.K.N., A.T.

Providing genotype and phenotype data: U.V., H.V., H.J.G Critical review of the manuscript: all authors

## Data and code availability

Developed scripts for the workflow are available on github (https://github.com/GenEpi-psych-UMG/Two_Step_Imputation). The data of the SHIP study cannot be made publically available due to the informed consent of the study participants, but it can be accessed through a data application form available at https://transfer.ship-med.uni-greifswald.de/ for researchers who meet the criteria for access to confidential data. The full results of the GWAS summary statistics are available on the ThyroidOmics Consortium website (http://www.thyroidomics.com).

## Consents and approvals

The study followed the recommendations of the Declaration of Helsinki. The medical ethics committee of the University of Greifswald approved the study protocol, and oral and written informed consents were obtained from each of the study participants.

## Conflicts of interest

The authors have no affiliation with any organization with a direct or indirect financial interest in the subject matter discussed in the manuscript. HJG received travel grants and speakers honoraria from Neuraxpharm, Servier, Indorsia and Janssen Cilag not related to the current project. HV received travel grants and speakers honoraria from Sanofi-Aventis not related to the current project.

