## Supplementary Information for "Removing array-specific batch effects in GWAS mega-analyses by applying a two-step imputation workflow reveals new associations for thyroid volume and goiter"

*Nasr MK et al.*

Supplementary Information

Supplementary Table 1. Distribution of the included variants for imputation against 1000G reference panel in the proposed two-step imputation with applied $R^2$ threshold 0.8 and 0.9 respectively. ... 14

### Supplementary Methods

#### Summary of the included cohorts

*SHIP*

The Study of Health in Pomerania (SHIP-START and SHIP-TREND) are parts of the Community Medicine Research network of the University of Greifswald, Germany, which are funded by the multiple institutions (grants no. 01ZZ9603, 01ZZ0103, 03Z1CN22, and 01ZZ0403), SHIP is a population-based project conducted in West Pomerania, a region in the northeast of Germany, consisting of two independent, prospectively collected cohorts (SHIP-START and SHIP-TREND), with an objective of assessing the prevalence and incidence of common population-based diseases and their associated risk factors[1]. A sample of the region's population who have German citizenship and main residency was randomly drawn and stratified by age and gender. Baseline examinations were carried out for SHIP-START from 1997 until 2001, resulting in a sample of 4,308 participants. While the baseline examinations for SHIP Trend were carried out between 2008 and 2012, with 4,420 individuals included.

*GANI_MED*

The research project "Greifswald Approach to Individualized Medicine" (GANI_MED) is a cohort with a focus on cardiovascular, cerebrovascular and metabolic diseases. The primary aim of this cohort is to increase the number of therapeutic strategies in personalized medicine approach[2]. Official patient recruitment started in 2011 with more than 4000 individuals included in the study.

#### Array panels included in the imputation workflow

*Affymetrix SNP 6.0*

The Affymetrix SNP 6.0 array contains more than 906,600 single nucleotide variants (SNVs) presented on 200 to 1,100 base pairs. SNVs are amplified using the Genome-Wide Human SNP Nsp/Sty Assay Kit 5.0/6.0, which is also validated for use for whole-genome sampling assay. This array was utilized for genotyping individuals from SHIP-START, comprising 4,070 individuals who were genotyped and subjected to quality control (QC) filtering.

*Affymetrix Axiom*

Axiom array was used for genotyping subgroup of 48 individuals from SHIP-START. This array features 560,000 SNVs available on the array, mostly available in HapMap. Two rounds of SNV selection were conducted with separate set of SNVs. This is considered to be of important biological value.

*Illumina Omni 2.5*

With more than 2.4 million SNVs per array, the Omni 2.5 delivers the most comprehensive coverage of SNVs among the included arrays in our project. This array was used for genotyping 986 individuals from SHIP-TREND.

*Illumina GSA*

The Infinium Global Screening Array (GSA) V3.0 is an advanced genotyping array which includes a multiethnic genome-wide backbone. GSA was used for genotyping 3,133 individuals from SHIP-TREND who are not overlapping with individuals genotyped with Omni 2.5 and included in our project.

*Illumina PsychArray*

The Illumina PsychArray array was designed with a focus on psychiatric disorders risks, covering more than 560,000 variants including tag SNVs from the HumanCore and Exome BeadChip, and other markers associated with common psychiatric disorders. 2,410 individuals from GANI_MED who were genotyped by this array were included in the project.

**Participant information and GWAS methodology for thyroid volume and goiter risk**

We included European ancestry participants from three genotyped SHIP cohorts (SHIP-START, SHIP-TREND and SHIP-TREND batch II). A total of 6,894 individuals, aged 20 to 83, have been included from for association testing. The GWAS was conducted twice for each investigated trait. One approach was conducted using the combined two-step imputed genotypes. Another one using conventionally imputed genotypes in each cohort separately, followed by meta-analysing the summary statistics of the three included cohorts.

All included individuals have a valid thyroid volume measurement determined by ultrasonography using an Ultrasound VST-Gateway (Diasonics, Santa Clara, USA) beside their quality controlled genotype data. To eliminate the confounding effect, we excluded individuals who are diagnosed with thyroid disorders or taking thyroid medication. Pregnant individuals were also excluded from the analysis. Goiter risk was identified as a dichotomous variable from the thyroid volume, all individuals with a thyroid volume above 25 and 18 ml for males and females respectively were identified with goiter risk. We used the log transformed values of the thyroid gland's volume for GWAS analysis. Both models were adjusted for age, sex, current smoking status, and body surface area (0.007184*(weight in kilograms$^{0.425}$) *(height in centimetres$^{0.725}$)) as covariates.

The statistical testing was performed using EPACTS software pipeline, using linear Wald testing for quantitative thyroid volume trait, and logistic Wald testing for the binary goiter risk trait[3]. For the meta-analysis of the single cohort GWAS summary statistics genomic control was applied to the individual GWAS result files if $\lambda_{GC} > 1$, and variants with an effective sample size < 75% of the total (maximum) sample size were excluded after the meta-analysis. Only autosomal variants with a minor allele frequency above 0.01 were included. The threshold for genome-wide significance was set at $5*10^{-8}$.

### Supplementary Figures

**Supplementary Figure 1. Genetic PCs (5-20) with their explained variance for conventional imputation (panel A) and two-step imputation (panel B)**

**A)**

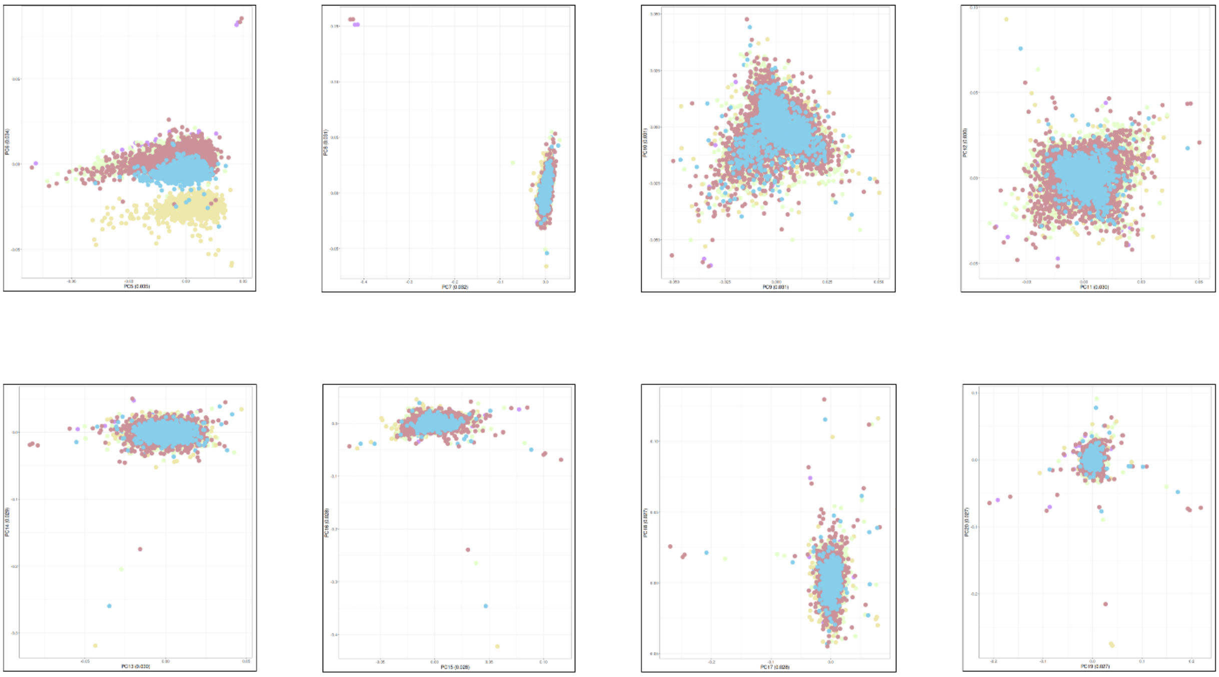

**B)**

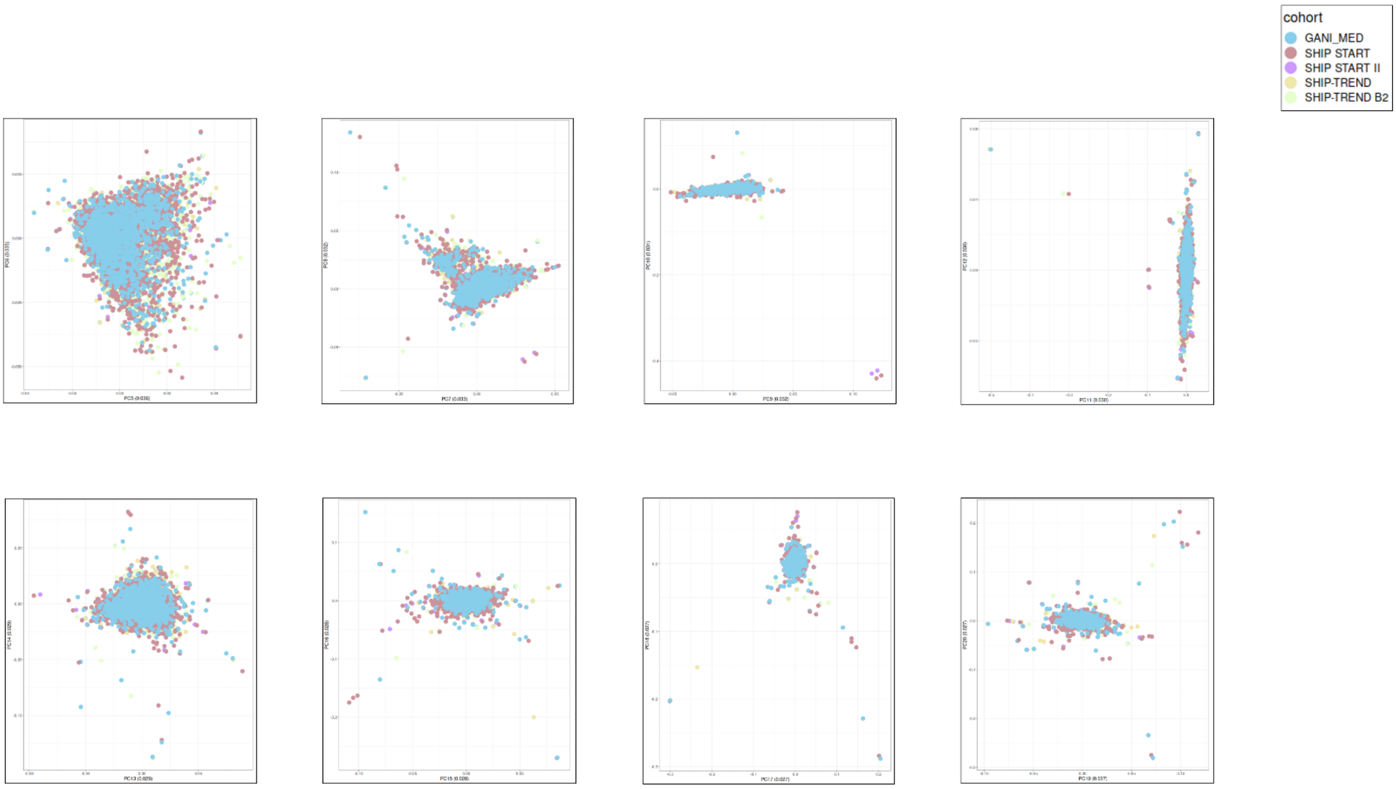

The samples are colored by the cohorts with their unique array type.

**Supplementary Figure 2. Genetic PCs (1-6) with their explained variance for two-step imputation using rare variants (MAF < 0.01)**

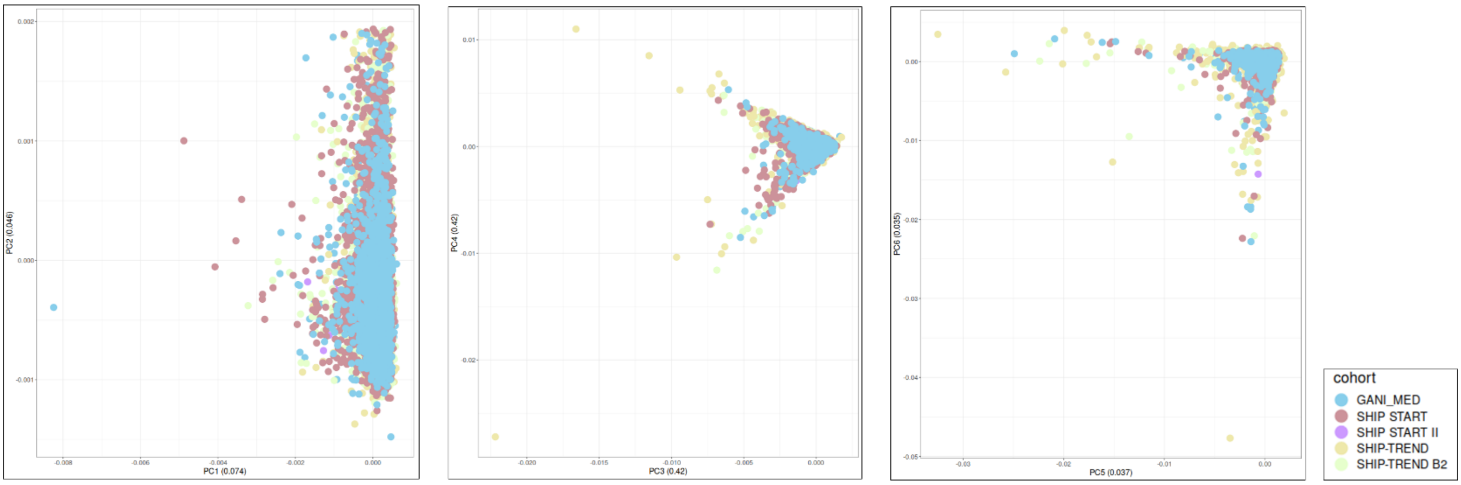

The samples are colored by the cohorts with their unique array type.

**Supplementary Figure 3. Genetic PCs (1-6) with their explained variance for conventional imputation using overlapped genotyped variants for imputation**

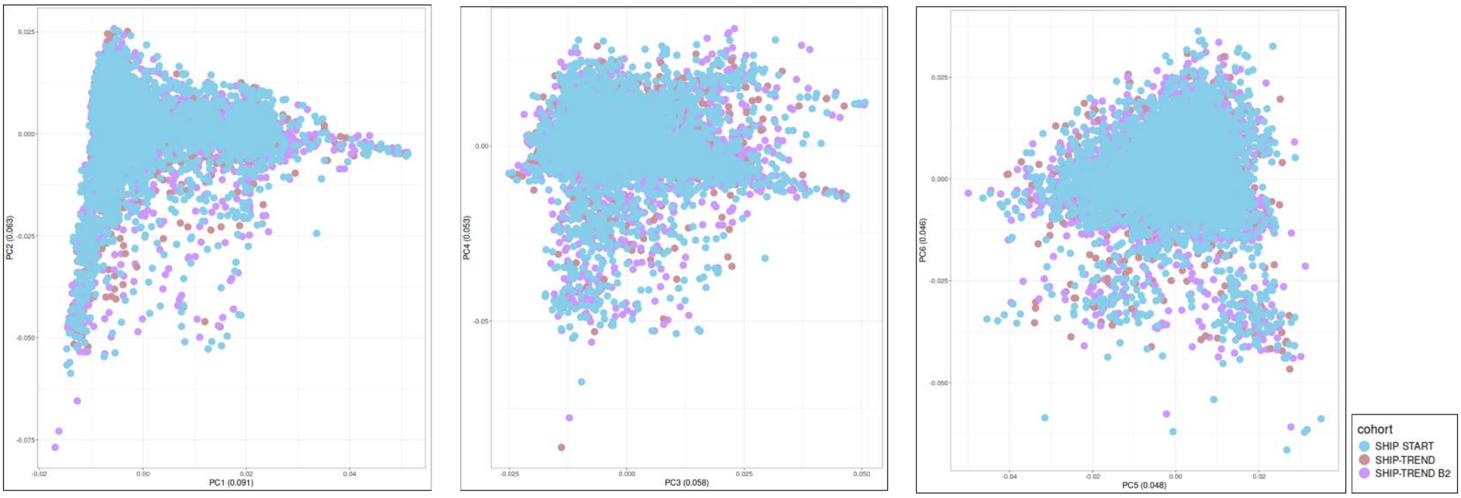

The samples are colored by the cohorts with their unique array type.

**Supplementary Figure 4. Median $R^2$ of the imputation outcomes of different approaches.**

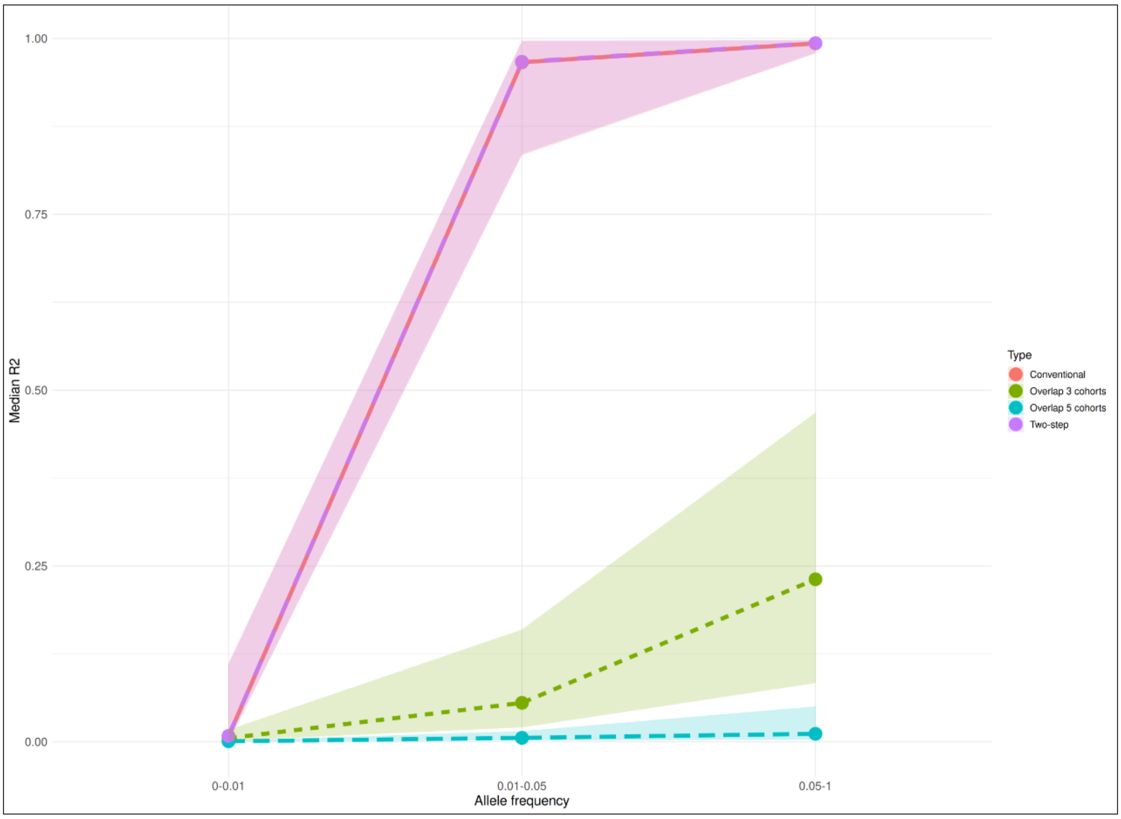

X axis represents allele frequency group ranging from 0.001 to monomorphic variants.

**Supplementary Figure 5. Imputation quality of the imputed variants.**

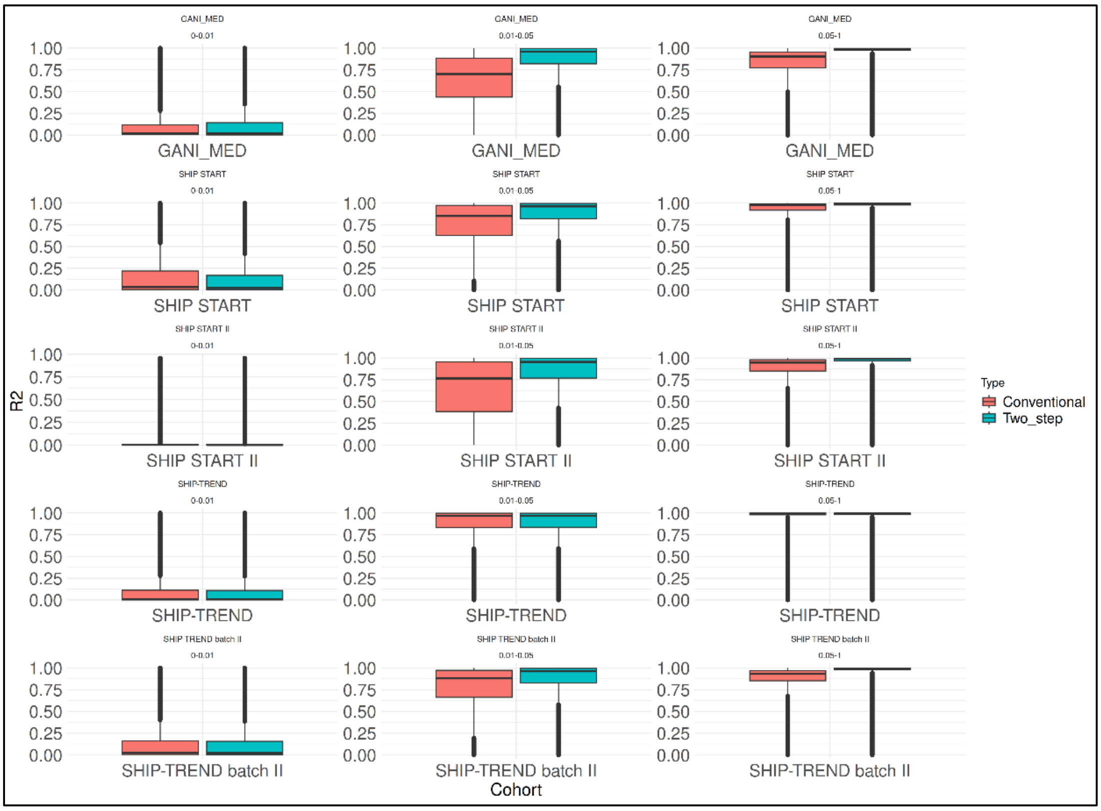

Boxplots grid of the imputation quality ($R^2$) strata of the imputed variants colored by the imputation type. Each column represents allele frequency group, and each row represents one of the included cohorts.

**Supplementary Figure 6. Genotype concordance**

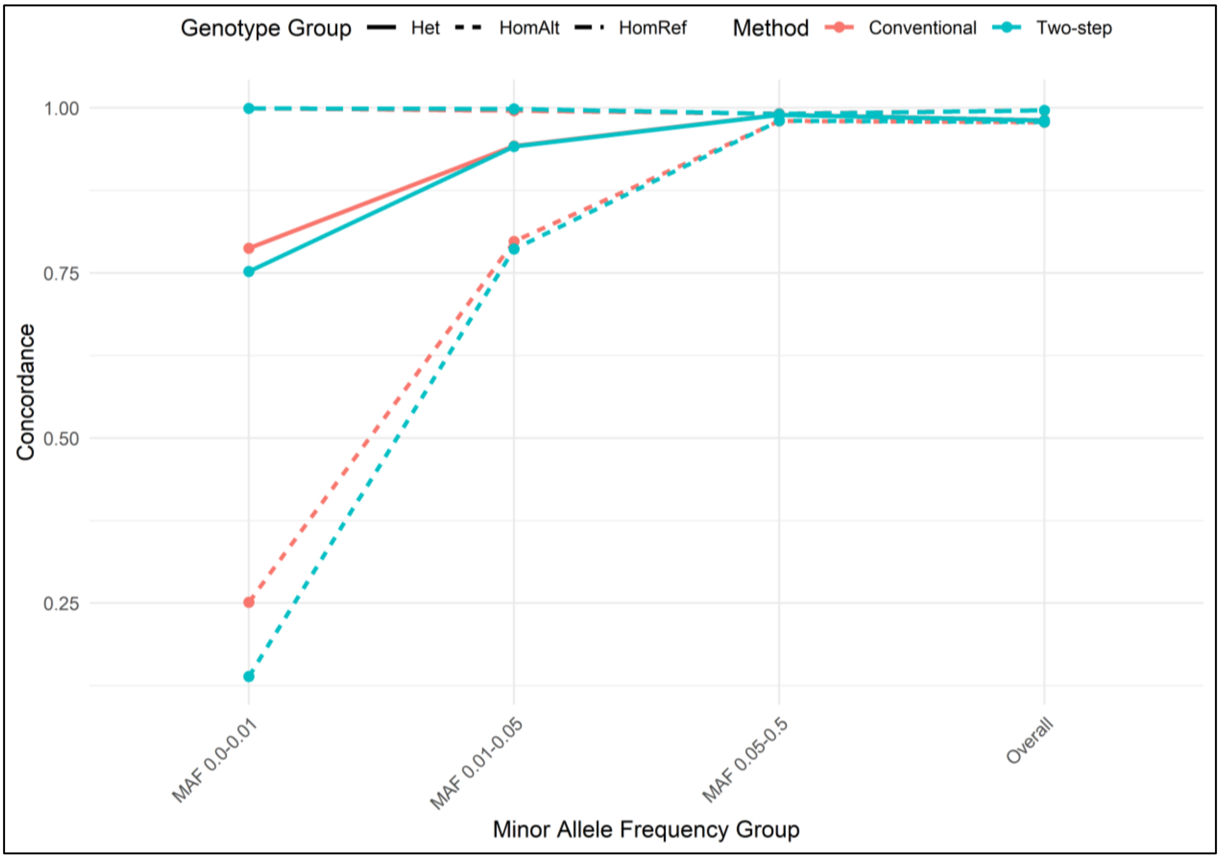

Concordance of the matching of the hard call imputed genotypes with sequenced data for homozygous reference (HomRef), homozygous alternative (HomAlt), and heterozygous (Het) calls, all variants followed by stratification by minor allele frequency (MAF) group of the imputed data.

**Supplementary Figure 7. Quantile-Quantile plots of the GWAS results**

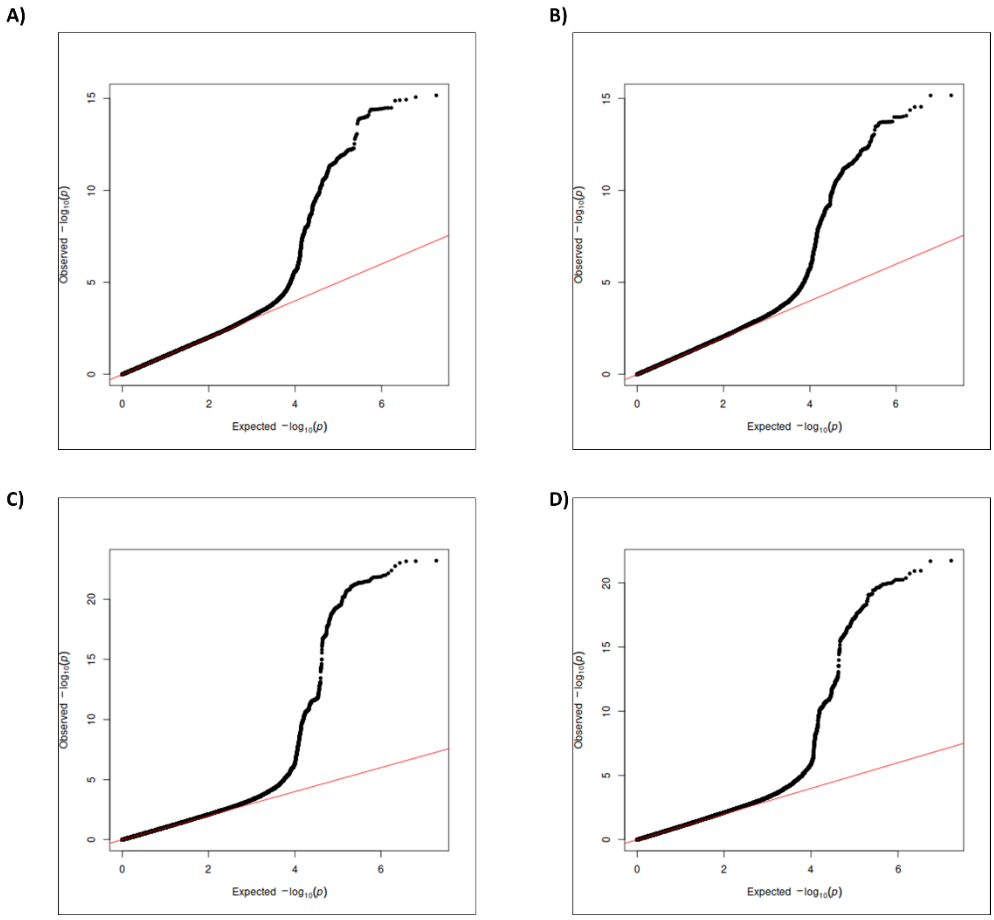

QQ plots for goiter risk using conventional imputation (panel A, $\Lambda_{GC}$ = 1.001), the combined two-step imputation (panel B, $\Lambda_{GC}$ = 1.030), and log thyroid volume using conventional imputation (panel C, $\Lambda_{GC}$ = 1.036) and the combined two-step imputation (panel D, $\Lambda_{GC}$ = 1.038)

**Supplementary Figure 8. Goiter risk GWAS summary statistics parameters**

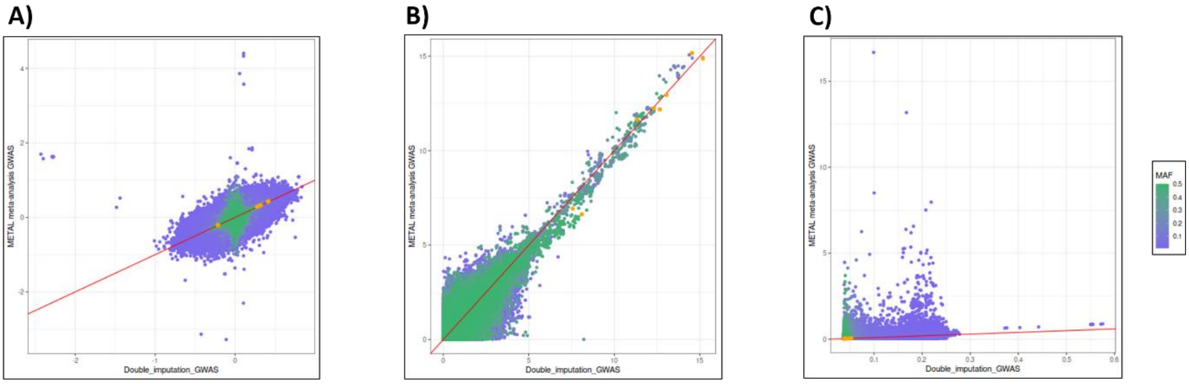

Scatter plots of effect estimates, -log 10 p-values and standard errors (panels A, B and C respectively) for two conducted GWAS approaches. Points are colored by minor allele frequency of the two-step imputed genotypes. Points colored in yellow represent the SNVs with significant association with goiter risk in both GWAS analysis

**Supplementary Figure 9. Log thyroid volume GWAS summary statistics parameters**

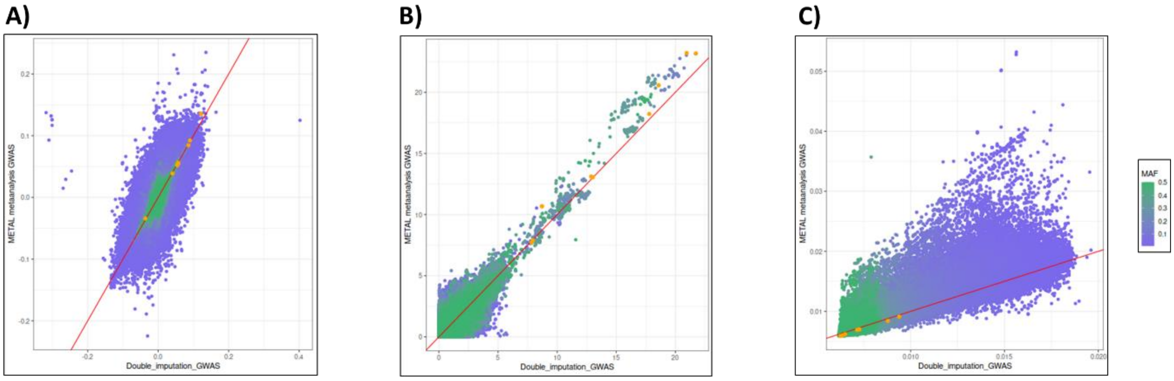

Scatter plots of effect estimates, -log 10 p-values and standard errors (panels A, B and C respectively) for two conducted GWAS approaches (x and y axis). Points are colored by minor allele frequency of the two-step imputed genotypes. Points colored in yellow represent the SNVs with significant association with log thyroid volume in both GWAS analysis

**Supplementary Figure 10. Genotyped versus sequenced allele frequencies**

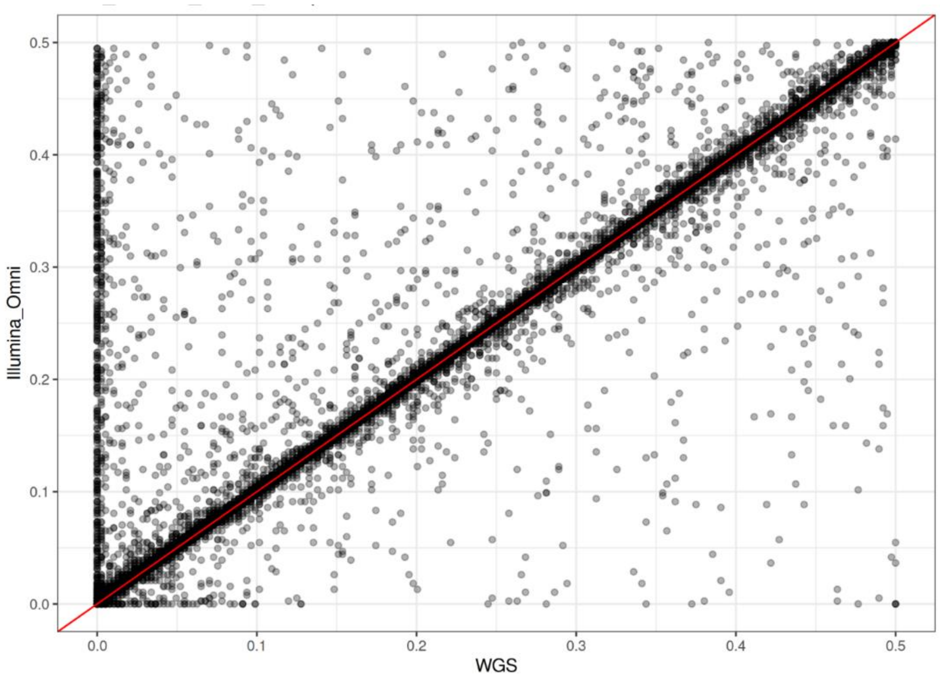

Minor allele frequencies in SHIP-TREND subgroup (n= 192) genotyped with Illumina Omni 2.5 (y axis) versus whole genome sequencing (x axis). Variants with missing genotypes were filtered out.

**Supplementary Figure 11. Genetic PCs (1-4) with the explained variance for two-step imputation upon using imputation quality threshold for intermediate imputation $R^2 > 0.8$**

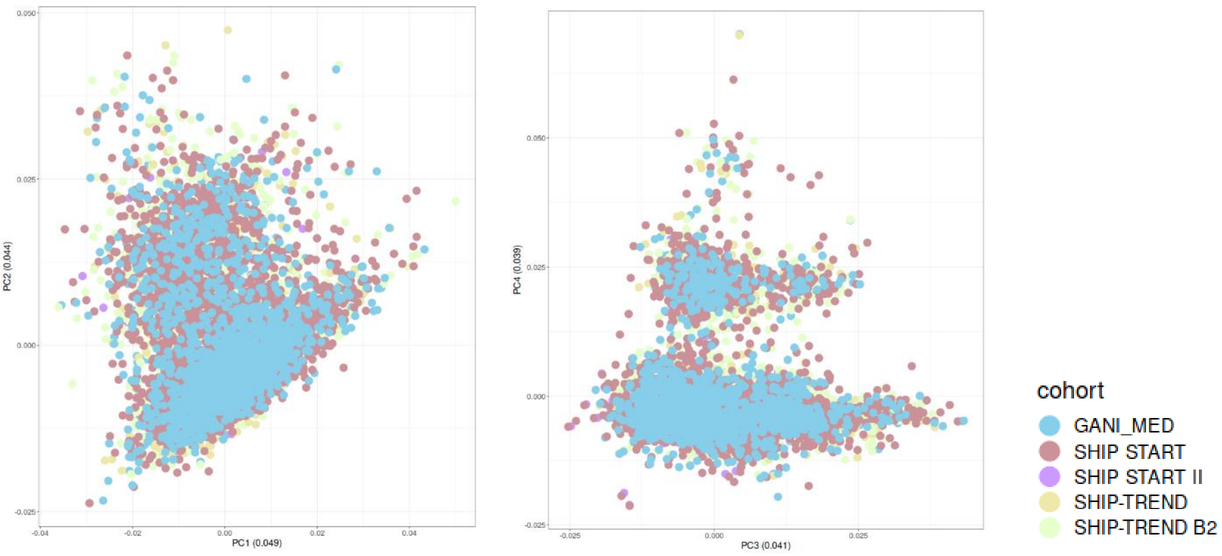

**Supplementary Figure 12. Minor allele frequency of SHIP-TREND imputation**

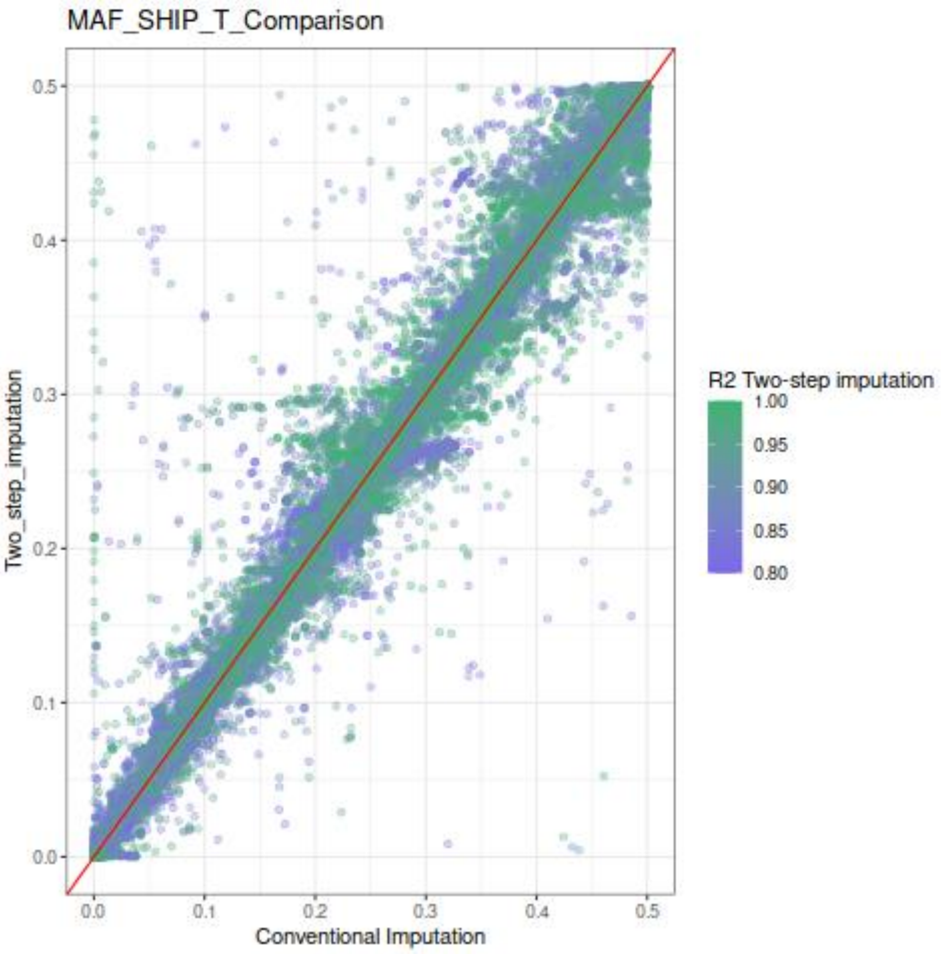

Scatter plot of the allele frequency between conventional (x axis) and two-step imputation (y axis) outcomes coloured by the imputation $R^2$ of the two-step imputation. Variants were selected by a minimum $R^2$ = 0.8.

### Supplementary Tables

**Supplementary Table 1. Distribution of the included variants for imputation against 1000G reference panel in the proposed two-step imputation with applied $R^2$ threshold 0.8 and 0.9 respectively.**

| chromosome | $R^2$ threshold = 0.8 | $R^2$ threshold = 0.9 | Overlap 5 cohorts | Overlap 3 cohorts |
|---|---|---|---|---|
| 1 | 161668 | 146001 | 935 | 4756 |
| 2 | 174018 | 155530 | 958 | 4684 |
| 3 | 146475 | 130690 | 904 | 3926 |
| 4 | 135205 | 121440 | 728 | 3387 |
| 5 | 130083 | 115862 | 728 | 3492 |
| 6 | 141011 | 124891 | 976 | 4025 |
| 7 | 115696 | 103837 | 736 | 3217 |
| 8 | 114410 | 101695 | 633 | 3003 |
| 9 | 94958 | 85149 | 569 | 2666 |
| 10 | 108586 | 97452 | 624 | 3121 |
| 11 | 104556 | 93927 | 566 | 2981 |
| 12 | 100130 | 89775 | 656 | 2909 |
| 13 | 76257 | 67913 | 501 | 2196 |
| 14 | 68389 | 62085 | 432 | 1940 |
| 15 | 63205 | 57674 | 383 | 1912 |
| 16 | 67940 | 62968 | 402 | 1970 |
| 17 | 56561 | 53446 | 338 | 1669 |
| 18 | 60575 | 55305 | 386 | 1797 |
| 19 | 42058 | 40105 | 227 | 1110 |
| 20 | 50755 | 46494 | 304 | 1617 |
| 21 | 28374 | 26104 | 175 | 879 |
| 22 | 30017 | 28608 | 104 | 834 |
| Total | 2070927 | 1866951 | 12265 | 58091 |

The last two columns show the distribution of the overlapped variants in all 5 cohorts and only 3 SHIP cohorts, respectively.

**Supplementary Table 2. Differences in median $R^2$ between conventional and two-step imputation approaches**

| Cohort | AF category | Median $R^2$ (conventional) | Median $R^2$ (Two-step) | Median MAF difference |
|---|---|---|---|---|
| GANI_MED | 0-0.001 | 0.01392 | 0.01094 | 0.00008 |
| | 0.001-0.01 | 0.2227 | 0.56329 | 0.00098 |
| | 0.01-0.02 | 0.63723 | 0.905425 | 0.00207 |
| | 0.02-0.03 | 0.71919 | 0.96703 | 0.00253 |
| | 0.03-0.04 | 0.76575 | 0.98345 | 0.00279 |
| | 0.04-0.05 | 0.79908 | 0.98827 | 0.00303 |
| | 0.05-1 | 0.90394 | 0.99093 | 0.00695 |
| SHIP-START | 0-0.001 | 0.02052 | 0.01386 | 0.00004 |
| | 0.001-0.01 | 0.4101 | 0.57362 | 0.00067 |
| | 0.01-0.02 | 0.7702 | 0.90913 | 0.00121 |
| | 0.02-0.03 | 0.86806 | 0.96962 | 0.0013 |
| | 0.03-0.04 | 0.92255 | 0.98555 | 0.00128 |
| | 0.04-0.05 | 0.94684 | 0.98974 | 0.00128 |
| | 0.05-1 | 0.97457 | 0.9921 | 0.00207 |
| SHIP-START II | 0-0.001 | 0.00076 | 0.00016 | 0.00004 |
| | 0.001-0.01 | 0.061155 | 0.10509 | 0.00172 |
| | 0.01-0.02 | 0.64082 | 0.89676 | 0.00465 |
| | 0.02-0.03 | 0.787305 | 0.96166 | 0.005 |
| | 0.03-0.04 | 0.86594 | 0.98326 | 0.00495 |
| | 0.04-0.05 | 0.90454 | 0.98996 | 0.00494 |
| | 0.05-1 | 0.94719 | 0.98935 | 0.00822 |
| SHIP-TREND | 0-0.001 | 0.0033 | 0.00373 | 0 |
| | 0.001-0.01 | 0.63978 | 0.63666 | 0.00016 |
| | 0.01-0.02 | 0.91488 | 0.91633 | 0.00017 |
| | 0.02-0.03 | 0.97447 | 0.97491 | 0.00012 |
| | 0.03-0.04 | 0.98762 | 0.98785 | 0.0001 |
| | 0.04-0.05 | 0.99217 | 0.99227 | 0.00008 |
| | 0.05-1 | 0.993 | 0.99336 | 0.00017 |
| SHIP-TREND B2 | 0-0.001 | 0.01563 | 0.01239 | 0.00006 |
| | 0.001-0.01 | 0.37792 | 0.57986 | 0.00078 |
| | 0.01-0.02 | 0.81587 | 0.91021 | 0.00124 |
| | 0.02-0.03 | 0.90292 | 0.96922 | 0.00132 |
| | 0.03-0.04 | 0.92814 | 0.98479 | 0.00139 |
| | 0.04-0.05 | 0.93044 | 0.9892 | 0.00154 |
| | 0.05-1 | 0.93135 | 0.99131 | 0.00443 |

Results of the included five cohorts are stratified by allele frequency (AF)

**Supplementary Table 3. Genotype concordance stratified by imputation quality.**

| Group | All variants | | $R^2 \geq 0.7$ | | $0.7 > R^2 \geq 0.3$ | | $R^2 < 0.3$ | |
|---|---|---|---|---|---|---|---|---|
| | Conventional | Two-step | Conventional | Two-step | Conventional | Two-step | Conventional | Two-step |
| **HomRef** | 0.9965 | 0.9962 | 0.9963 | 0.9962 | 0.9952 | 0.995 | 0.9922 | 0.9915 |
| **Het** | 0.982 | 0.981 | 0.987 | 0.9866 | 0.6388 | 0.6347 | 0.9231 | 0.923 |
| **HomAlt** | 0.9785 | 0.9787 | 0.9815 | 0.9818 | 0.5423 | 0.5434 | 0.9333 | 0.936 |

Genotype concordance (number of matching genotypes/total number of genotypes) of the hard call imputed genotypes with sequenced data for homozygous reference (HomRef), homozygous alternative (HomAlt), and heterozygous (Het) calls, all variants followed by stratification by imputation quality $R^2$.
